# Hospital Acquired Pneumonia Prevention (HAPPEN) Study: Statistical Analysis Plan

**DOI:** 10.1101/2025.08.14.25333719

**Authors:** Nicole M White, Allen Cheng, Andrew Stewardson, Philip L Russo, Brett Mitchell

## Abstract

The Hospital-acquired Pneumonia Prevention (HAPPEN) study is a multi-site trial evaluating the clinical effectiveness of improving oral care among adult patients to reduce the incidence of non-ventilator, hospital-acquired pneumonia (HAP). The HAPPEN study design is a stepped-wedge cluster randomised trial across three public hospitals in Australia. This document is the Statistical Analysis Plan for evaluating primary and secondary effectiveness outcomes. The trial was preregistered on the Australian and New Zealand Clinical Trials registry (ACTRN12624000187549) and is funded by the Medical Research Future Fund (MRF2022645). A copy of the study protocol and a signed version of the Statistical Analysis Plan are available on request from the corresponding author (BM).

## Administrative information

### 1.1 STUDY IDENTIFIERS

- Protocol: V1.3
- Medical Research Future Fund: MRF2022645
- ANZCTR: ACTRN12624000187549

### 1.2 REVISION HISTORY

| Version | Date | Changes made to document | Authors |
| --- | --- | --- | --- |
| 1.1 |  | First version | NW, BM. |
| 1.2 |  | Revised based on initial CI feedback | NW, BM, AS, AC |
| 1.3 | 7<sup>th</sup> August 2025<br>(approved version) | Revised following feedback received at CI meeting | NW, BM, AS, AC, PR |

### 1.3 CONTRIBUTORS TO THE STATISTICAL ANALYSIS PLAN

#### 1.3.1 ROLES AND RESPONSIBILITIES

1.3 CONTRIBUTORS TO THE STATISTICAL ANALYSIS PLAN 1.3.1 ROLES AND RESPONSIBILITIES
| Names and OCRID | Affiliation | Role on study | SAP contribution |
| --- | --- | --- | --- |
| Nicole White | Queensland University of Technology | Chief Investigator | Development |
| Brett Mitchell | Avondale University and Central Coast Local Health District | Chief Investigator | Critical review and input |
| Allen Cheng | Monash Health and Monash University | Chief Investigator | Critical review and input |
| Andrew Stewardson | Alfred Health and Monash University | Chief Investigator | Critical review and input |
| Philip Russo | Cabrini Health and Monash University | Chief Investigator | Critical review and input |

## Study overview

### Synopsis

The Hospital-acquired Pneumonia prevention (HAPPEN) study is a multi-site trial evaluating the clinical effectiveness of improving oral care among adult patients to reduce the incidence of non-ventilator, hospital-acquired pneumonia (HAP).

The trial will evaluate the change in HAP incidence from improving the frequency and quality of oral care among patients admitted to participating hospital wards, compared with the current standard of oral care.

### Study design

The HAPPEN study design is a stepped-wedge cluster randomised trial across three public hospitals in Australia (Figure 1). The chosen study design allows patients admitted to the participating wards the opportunity to benefit from the intervention.

**Figure 1:**
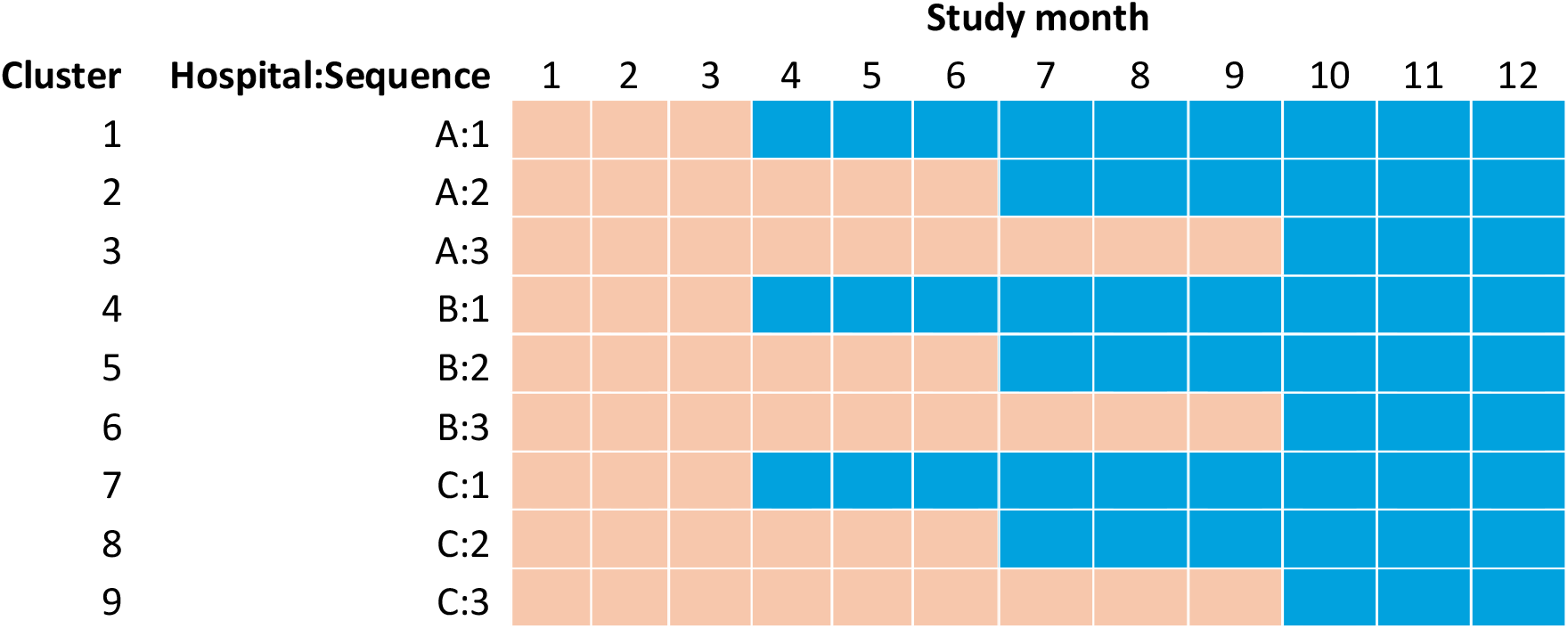
Study design.

The study design consists of nine clusters, where a single ward is defined as a cluster. Three wards from each participating hospital will be enrolled in the study. Wards will be randomised to one of three treatment sequences, which will determine when they transition from the control condition (current standard of oral care) to the intervention condition (oral care bundle).

All clusters will start in the control condition for the first three months of the study. One ward from each hospital will then transition to the intervention condition every 3 months until all wards have been exposed to the intervention. The study timeframe is 12 months, which corresponds to an intervention exposure time of 3 to 9 months, based on a cluster’s allocated treatment sequence. The length of each period is 1 month.

No transition phase is assumed, meaning that intervention exposure will begin on the same day a cluster transitions to the intervention condition.

## Study objectives

### Primary objective

To evaluate the clinical effectiveness of improving oral care among enrolled patients admitted to study wards for 48 hours or more on new cases of non-ventilator-associated hospital-acquired pneumonia.

The hypothesis to be tested is that improving the frequency and quality of oral care will reduce HAP incidence across study wards, compared with the current standard of oral care in study hospitals.

### Secondary objectives

To evaluate the clinical effectiveness of the intervention on the incidence of other respiratory and oral infections in study wards. This objective is motivated by the hypothesis that improved oral care frequency and quality may reduce the transmission of other pathogens found in hospitals.

A health economic evaluation of the intervention will also be conducted as a secondary objective, using results generated by this Statistical Analysis Plan. This evaluation will be detailed in a separate Health Economic Analysis Plan.

## Study population

The study population is defined in terms of eligible hospitals and eligible patients.

Eligible hospitals will be Group A public acute hospitals^1^ located in Australia which have an intensive care unit. In addition to these criteria, hospitals must have at least 10 adult wards and an average of 50,000 admissions per year. These additional criteria are to provide a sufficient sampling frame for the chosen study design and to meet sample size requirements.

Eligible patients are adults aged 18 years or older who have been admitted to a study ward for 48 hours or more (Figure 2). For patients who meet the inclusion criteria, confirmed infections will be excluded if acquired on the day of study ward admission or within the first 48 hours of admission.

**Figure 2:**
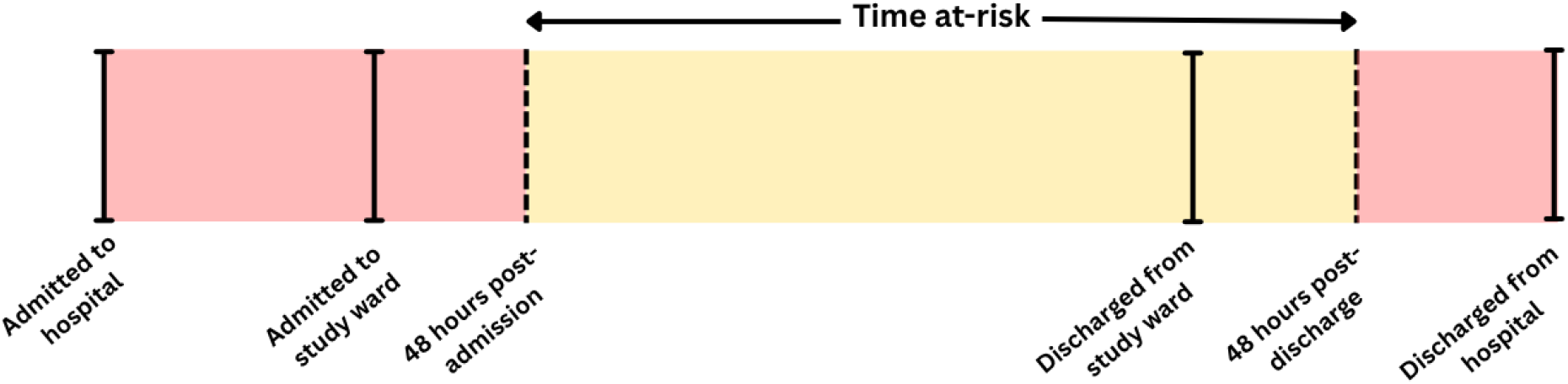
Time at-risk definition for confirmed infections.

Eligible patients admitted to each ward (cluster) during a single period will receive either the control or intervention condition. As some patients may remain on the ward for multiple periods, there is a chance that they are exposed to both control and intervention conditions. Attribution rules for confirmed infections are outlined in the next Section.

## Outcomes

### Primary outcome

The primary outcome is non-ventilator-associated hospital-acquired pneumonia (HAP). The primary outcome definition is the European Centre for Disease Control definition of HAP (1).

Primary outcome data will be collected for all patients admitted to study wards using a REDCap algorithm to identify new HAP cases. The algorithm has been previously tested and validated in other studies (2, 3). An interrater reliability assessment will be independently undertaken to validate the algorithm’s accuracy for detecting HAP.

## Secondary outcomes

The incidence of upper and lower respiratory infections combined, and EENT Oral infection (EENT-O) will be analysed as secondary outcomes. These secondary outcomes have been selected to evaluate given the intervention’s potential to reduce the risk of patients acquiring other respiratory pathogens.

The observational unit for primary and secondary outcomes is defined as a single patient admission. Outcomes will be assessed at the end of a patient’s admission, to determine if the patient acquired an infection and if so, when the infection occurred.

The following rules will be applied to attribute new infections found by the algorithm to study wards and study conditions (Figure 3):

**Figure 3:**
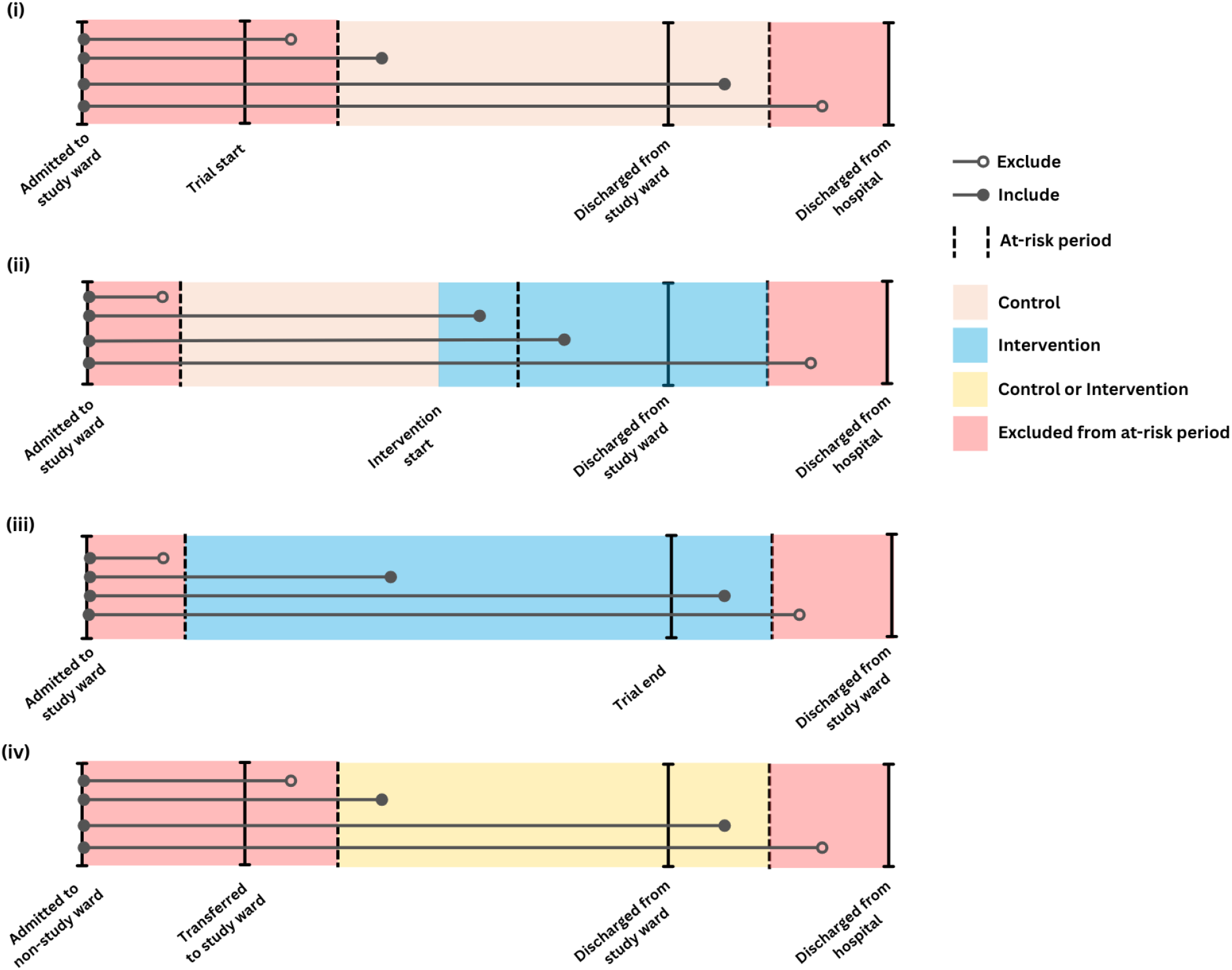
Attribution rules for confirmed infections - primary and secondary outcomes.

i. If a patient is admitted to a study ward before the trial start date and the date of infection onset is less than 2 days after the trial start date, the infection will be excluded.
ii. If a patient is admitted to a ward before the start of intervention exposure and acquires an infection on the same ward after being exposed to the intervention,
  a. The infection will be attributed to the control condition if the date of onset is within two days of the ward’s intervention start date.
  b. The infection will be attributed to the intervention condition if the date of onset is more than 2 days after the ward’s intervention start date.
iii. If a patient is admitted to a study ward under the intervention condition, and acquires an infection in the same ward after the trial end date:
  a. The infection will be attributed to the intervention condition if the date of onset is within two days after the study end date.
  b. The infection will be excluded if the date of onset is more than 2 days after the trial end date.
iv. If a patient is transferred between wards and acquires an infection:
  a. The infection will be attributed to the ward the patient was transferred from if date of onset is less than 2 days after transfer.
  b. The infection will be attributed to the ward the patient was transferred to if date of onset is 2 or more days after transfer.

These rules have been encoded into the REDCap algorithm and will be rechecked as part of data cleaning before analysis.

## Intervention

The intervention combines an oral care component with patient and staff education, and audit and feedback to ward nursing staff. The oral care component will provide all eligible patients with an oral care protocol, aiming for twice-daily use, increasing to 4-hourly for participants who are nil per by mouth or require assistance with oral care. Participating hospitals will be provided with oral care packs for eligible patients to implement the intervention.

All clusters will receive the intervention based on randomised treatment allocations in Figure 1. The intervention will be delivered to all eligible patients in a cluster once the cluster switches from the control condition to intervention condition.

## Randomisation and blinding

The trial statistician completed randomisation on 22 May 2024, after all three hospitals had been recruited to the trial and before the start of the control period (3 June 2024). Ward (cluster) randomisation to treatment sequences (1, 2 or 3) was performed by hospital using a set seed in the R statistical software. Clusters will be labelled 1 to 9 in the final dataset, as shown in Figure 1, for use in analysis and reporting. For hospital-level analyses, individual hospitals will be labelled as A, B or C, per Figure 1.

The trial statistician, principal investigator and research coordinator will be unblinded to the treatment sequences and are not involved in data collection.

Study team members responsible for collecting outcome data were blinded to allocated treatment sequences and, therefore, did not know when wards transitioned from the control period to the intervention period or how long each ward had been exposed to the intervention.

It was not possible to blind patients or ward nursing staff following intervention exposure, given their involvement in complying with the intervention and the provision of monthly feedback to staff about intervention compliance and the number of new HAP cases. Wards will be informed of their intervention start date four weeks prior.

## Sample size

Sample size calculations align with the project protocol, which are based on the primary outcome using the study design shown in Figure 1. Calculations were powered to test for a statistically significant reduction in the primary outcome associated with intervention exposure, assuming a two-sided alternative hypothesis and a 5% level of statistical significance.

HAP incidence during the control period was assumed to be 7 infections per 100 admissions. This estimate was informed by published data collected on similar high-risk adult populations (4, 5). Calculations were based on a fixed sample size per cluster-period (125 ward admissions per month), with a coefficient of variation equal to 0.5 applied to account for possible variation in cluster sizes (6). Initial calculations treated the primary outcome as a proportion (number of HAP cases divided number of patients at-risk) assuming an exchangeable correlation structure and intraclass correlation coefficient (ICC) set to 0.02. Our chosen ICC is conservative and reflect the assumption that patient outcomes are relatively independent; if the observed ICC is higher than 0.02, the expected power will increase, holding all other parameters constant. We also considered calculations where the primary outcome was treated as a count (confirmed HAP cases per 100 admissions), to assess sensitivity to distributional assumptions.

As a binary outcome expressed as a proportion, a minimum sample size of 125 per cluster-period returns 80% power to detect a 30% decrease in HAP incidence from 0.07 (7%) to 0.049 (4.9%) associated with the intervention. The impact of distributional assumptions was minimal - when modelled as a count, the same sample size is sufficient to detect a 31% decrease in HAP incidence from 7 to 4.8 cases per 100 admissions with 81% power. Both calculations correspond to a total trial sample size of 13,500 patients.

## Statistical analysis

### General principles

This plan outlines the analysis of the stated primary and secondary objectives. Additional objectives (e.g., health economic evaluation) will be detailed in a separate analysis plan.

All statistical analyses and graphics will be completed in R version 4.1.1 or higher. R packages used for analysis will be reported in a software statement in the final analysis report. Analysis code will be made publicly available on GitHub at https://github.com/nicolemwhite/HAPPEN.

The research coordinator will complete data validation and cleaning. The final dataset will be sent to the trial statistician in.csv or.xlsx format with a completed data dictionary. Further data processing by the trial statistician will include the merging of data files by unique identifier(s) (where applicable) and checking of date-time variables against control and intervention period definitions as per the study protocol. Systematic data quality checks (e.g., negative numbers of cases, implausible dates, duplicate records) will be run to identify potential data entry errors for resolution with the research coordinator before starting analysis. No interim analyses will be completed as no negative study effects are anticipated.

Analyses will be reported in accordance with the Consolidated Standards of Reporting Trials (CONSORT) extension for stepped wedge cluster randomised trials (https://www.bmj.com/content/363/bmj.k1614). A completed checklist will be provided as a supplementary file with the results manuscript.

### Summary of changes

As a tertiary outcome, primary and secondary infection outcomes will be analysed as a composite outcome. The composite outcomes will be defined as one or more of HAP, lower/upper respiratory infection, EENT-O.

Monthly audit data used for staff feedback on intervention fidelity will be collected over the trial. The data collected will report on the proportion of people who had oral care performed on the day the audit was completed.

### Sensitivity analysis

The main analysis of primary and secondary infection outcomes assumes a common and constant effect for intervention exposure. Resulting estimates therefore represent the average within-ward change in incidence from intervention exposure, compared with the current standard of oral care.

Sensitivity analyses will explore different assumptions about the effectiveness of the intervention:

i. Differences in intervention effectiveness by hospital. A per-hospital intervention effect will be added to primary and secondary infection models to test for differences in intervention effectiveness between the three study hospitals. The null hypothesis is that the estimated effect of the intervention is the same across all hospitals.
ii. Intervention effectiveness over time. Intervention exposure will be defined as time since a cluster’s transition from control to intervention conditions, in place of a binary fixed effect.
iii. Assessing the relative contribution of individual clusters. We will conduct a leave-one-cluster-out analysis and assess changes to estimated intervention effectiveness.
iv. HAP attribution to Control vs Intervention conditions. Since the intervention will only be trialled in 3 wards per hospital, it is possible that eligible patients are transferred between non-study and/or study wards and acquire HAP within 48 hours before or after transfer. To address this possibility, we will conduct a sensitivity analysis that attributes HAP cases to the control or intervention condition based on the recorded date of HAP onset (i.e. 48-hour washout period between control and intervention not applied).
v. Per-protocol analysis. In the event of study ward(s) cannot be accessed during the trial (e.g., closed due to an outbreak), we will re-run primary and secondary outcome model with affected cluster-periods excluded.

Sensitivity analyses are intended to be exploratory and reported after the main analysis of primary and secondary infection outcomes.

### Blind review

A blinded review is planned (except for the study Statistician and Lead Chief Investigator) until there is agreement of the final presentation of results.

### Data sets to be analysed

We will use an intention-to-treat analysis approach to analysis. A per-protocol analysis will be conducted if there are major deviations to the trial design, e.g., if the intervention did not start at the planned time, if an outbreak occurs on a study ward that prevents the intervention from being used.

### Subject disposition

A CONSORT flowchart will be used to summarise the number of eligible admissions and exclusions. Flowchart information will align with guidance provided by the cluster randomised trial extension to the CONSORT reporting checklist (7).

### Patient characteristics and baseline comparisons

Cohort characteristics will be summarised descriptively overall and by hospital, stratified by study condition (control, intervention). Other descriptive statistics will be presented in tabular format.

Continuous variables will be summarised as means with standard deviations or medians with lower and upper quartiles. Categorical variables will be summarised as frequencies (numerator and denominator) with percentages. Reported denominators will reflect the presence of any missing data. All patient characteristics will be reported as collected without transformation. There will be no hypothesis testing of differences in baseline characteristics between hospitals or study periods.

For all infections analysed, we will report total cases by study condition and infection type.

Frequencies will be complemented by unadjusted (except for clustering) incidence rates, reported as estimates with 95% confidence intervals.

### Primary outcome analysis

The primary outcome will be modelled at the cluster-level and the admission-level. Cluster-level analysis will model the number of new HAP events per 100 at-risk admissions, Admission-level analysis will model the cumulative incidence of HAP.

Generalized linear mixed models (GLMM) will analyse the within-ward change in the primary outcome associated with intervention exposure. GLMMs will assume logit and log link functions for cluster-level analyses to associate expected outcomes with fixed and random effects. A complementary log-log link function will be used to model cumulative incidence at the admission level.

The fixed effects of interest are intervention exposure and period. Each period will be defined as 1 month and represented in the GLMM as a categorical fixed effect. A linear effect for period will also be tested and reported if the categorical fixed effect specification does not converge. Intervention exposure will be defined as a categorical variable (no = 0, yes = 1). Intervention exposure will be coded as 0 for the entire control condition, then switched to 1 at the start of the intervention condition. This specification assumes a constant effect of the intervention. A random effect per ward will be specified to account for within-cluster correlation and differences in control HAP incidence.

GLMM results will be reported for all fixed effects as odds ratios or incidence risk ratios with 95% confidence intervals. Confidence intervals will use model-based standard errors with and without a degrees-of-freedom correction based on the number of clusters (8). Model-based estimates will be reported along with changes in absolute and relative risk, which will be estimated by bootstrapping.

Sensitivity analyses will consider the relative contribution of study hospital to estimated intervention effectiveness, and the specification of fixed effects to represent intervention exposure. Hospital-level risk contributions with intervention effectiveness will be examined by adding a random intervention effect per hospital. In place of a binary fixed effect, intervention exposure will be modelled in months to examine evidence of changes in intervention effectiveness over time.

Based on data collection instruments, we do not anticipate any missing data for the primary outcome. In the event of missing outcome data, these will be reported in the CONSORT study flowchart.

### Secondary outcome analysis

Respiratory tract (lower and upper combined) and EENT-O infections will be analysed using GLMMs (Table 1). Given the likelihood of multiple infections per admission, outcomes will be modelled as:

**Table 1:** Summary of primary, secondary, and tertiary outcome analysis.

| Study descriptor | Primary outcome | Secondary outcomes | Tertiary outcomes |
| --- | --- | --- | --- |
| Population | Patients aged 18 years or over admitted to trial study wards |  |  |
| Intervention | Provision of oral care protocol for eligible patients, combined with patient and staff education, and audit and feedback to ward nursing staff |  |  |
| Comparator | Current oral care standards in eligible hospitals |  |  |
| Outcome | HAP incidence<br>- Proportion of admissions<br>- Count of total cases per 100 admissions | Combined lower and upper respiratory tract infection incidence.<br>EENT-O incidence.<br>- The proportion of patients who acquire one or more infections during their admission<br>- Total infection per 100 admissions | Total infection incidence (HAP + Respiratory tract + EENT-O) |
| Statistical approach | GLMM | GLMM | GLMM<br>Intervention fidelity:<br>Descriptive analysis of oral care audit data |

Cluster-level

1. The proportion of patients who acquire one or more infections during their admission, and
2. The total number of infections per 100 at-risk admissions. Patient-level
3. The cumulative incidence of infection from admission to discharge from the study ward.

A binomial response with a logit link will be assumed for (1); (2) will be modelled using a Poisson distribution with a log link function and log person-time as an offset. As per the primary outcome, models will include a random intercept per ward and categorical fixed effects for intervention exposure and period.

The number of lower respiratory and upper respiratory infections will be reported descriptively to provide information about their relative contributions to their combined incidence.

### Tertiary outcome analysis

The total number of infections (HAP + Respiratory tract + EENT-O) will be analysed in terms of:

1. The proportion of patients who acquire one or more infections during their admission, and
2. The total number of infections per 100 admissions (Table 1).
3. The cumulative incidence of infection from admission to discharge from the study ward.

Intervention fidelity will be described descriptively, using monthly audit results on oral care. The outcome to be analysed is the proportion of eligible patients who had oral care performed.

Tertiary outcome results will be presented graphically to compare trends in composite infection outcomes and intervention fidelity. Associations between infections and fidelity will not be tested.

Key details for primary, secondary, and tertiary analyses are summarised in Table 1.

## Data Availability

All data produced in the present work are contained in the manuscript

## List of abbreviations

EENT-O: Eye, ear, nose, throat infections, oral.
GLMM: Generalised Linear Mixed Model
HAP: Non-ventilator, hospital-acquired pneumonia
HAPPEN study: The Hospital-acquired Pneumonia prevention study
ICC: Intraclass correlation coefficient

## Footnotes

1 https://www.aihw.gov.au/reports/hospitals/australian-hospital-peer-groups/formats

